# Cardioprotective Effects of Sodium-Glucose Cotransporter-2 Inhibitors in Patients undergoing Ventricular Tachycardia (VT) Ablation: A Propensity-Matched Cohort Study

**DOI:** 10.1101/2025.10.02.25337206

**Authors:** Muhammad Raffey Shabbir, Omer Adeeb, Ahmad Baig, Muhammad Taimur, Maria Shahzad, Ayesha Majid, Meenal Sikander, Mahrukh Imtiaz, Param P. Sharma, Muhammad Osama

## Abstract

**Introduction:** Cardiovascular disease is one of the leading causes of death in patients with type 2 diabetes mellitus (T2DM). Sodium-glucose cotransporter-2 inhibitors (SGLT2i) have proven to be beneficial in improving cardiovascular outcomes and reducing all-cause mortality in patients with T2DM. We aimed to investigate the effect SGLT2i use on cardiovascular outcomes inpatients with T2DM who underwent VT ablation.

**Methods:** A retrospective cohort study was conducted using the TriNetX US Collaborative Network, a federated network of healthcare organizations across the United States. Adults (aged 18–80 years) with T2DM who underwent catheter ablation for ventricular tachycardia were included. Patients were stratified based on exposure to SGLT2i. Propensity score matching (1:1) was used to balance baseline characteristics. Outcomes were assessed within 3 years following the index ablation procedure. Patients with a recorded occurrence of the outcome prior to the index event were excluded from each respective outcome analysis. Kaplan-Meier analysis and log-rank tests were used for statistical comparisons with significance set at p<0.05.

**Results:** SGLT2 inhibitor non-users exhibited significantly higher hazard ratios (HR) for various adverse outcomes. The HR for all-cause mortality was 1.422 (95% CI: 1.279–1.581), while the HR for cardiac arrest was 1.409 (95% CI: 1.135–1.750). Additionally, the HR for post-ablation cardioversion was 1.188 (95% CI: 1.042–1.355), and the utilization of amiodarone after ablation had an HR of 1.240 (95% CI: 1.106–1.391). In contrast, the hazard ratios for redo ablation (1.039, 95% CI: 0.956–1.128), visits for ICD adjustments (0.916, 95% CI: 0.766–1.096), post-ablation use of any class of antiarrhythmics (1.139, 95% CI: 0.906–1.431), and lidocaine (0.911, 95% CI: 0.775–1.070) were less definitive.

**Conclusion:** SGLT2i non-user group was associated with significantly higher risks of several adverse outcomes following ablation, including a 42% increase in all-cause mortality and a 41% increase in cardiac arrest. Non-users also had higher rates of post-ablation cardioversion and amiodarone use. However, no significant differences were found in redo ablation, ICD adjustments, or the use of other antiarrhythmics. These findings suggest a potential protective role of SGLT2 inhibitors in selective cardiovascular outcomes. Further studies are warranted to confirm these associations and investigate the underlying mechanisms.

## Introduction

Ventricular tachycardia (VT) is a wide complex tachycardia, defined as 3 or more consecutive beats at a rate of 100 per minute, arising from the ventricles (1). VT is a potentially life-threatening arrhythmia that often occurs in the setting of structural heart disease, particularly ischemic and non-ischemic cardiomyopathy. Symptoms can range from being well-tolerated to causing severe hemodynamic instability or sudden cardiac death (SCD) (2). Catheter ablation has been proven as an effective treatment option for patients with drug-refractory VT and in patients with increased frequency of implantable cardioverter-defibrillator shocks (3) (4). However, despite its effectiveness, a substantial proportion of patients experience VT recurrence post-procedure (5). This emphasizes the importance of identifying factors that influence long-term outcomes. SGLT2i, initially developed for glucose lowering in type 2 diabetes mellitus, have demonstrated cardiovascular benefits beyond glycemic control(6). Landmark trials including EMPA-REG OUTCOME, CANVAS and DECLARE-TIMI 58 have consistently shown reductions in major adverse cardiovascular events, heart failure hospitalizations, and cardiovascular mortality in both diabetic and non-diabetic individuals (7)(8))(9). Several studies have shown that in patients with T2DM, treatment with SGLT2i appears to be independently associated with a significant reduction in the risk of recurrent atrial arrhythmias after catheter ablation (10)(11). The specific impact of SGLT2i on outcomes following VT ablation remains unexplored. Prior studies have reported associations between use of SGLT2i and a reduction in frequency of arrhythmia episodes, fewer ICD-related interventions, and improved clinical outcomes, including lower risks of hospitalization and mortality (12)(13). However, larger and long-term clinical and observational studies are still needed to investigate effects of SGLT2 inhibitors in this regard. This study aims to fill this gap by comparing post-ablation outcomes between SGLT2i users and non-users.

## Methods

### Data Source and Study Design

We used the US Collaborative Network Healthcare Organizations (HCOs) in the TriNetX Research Network. The available data included information about the demographics, diagnoses (based on the International Classification of Diseases, Tenth Revision, Clinical Modification, ICD-10-CM codes), procedures (coded in The International Classification of Diseases, Tenth Revision, Procedure Coding System, ICD-10-PCS or Current Procedural Terminology, CPT), medication (coded in Veterans Affairs National Formulary), laboratory tests (coded in Logical Observation Identifiers Names and Codes, LOINC), genomics (coded in Human Genome Variation Society, HGVS), and healthcare utilization. The HCOs were hospitals, primary-care units, or specialists, providing data from uninsured or insured patients.

In this study, we used the US Collaborative Network in TriNetX to build a cohort of adults (aged 18–80 years) with Type 2 Diabetes Mellitus who underwent catheter ablation for ventricular tachycardia. Patients were stratified based on exposure to SGLT2i. We identified around 93 million participants to include in the study.

#### Ethics statement

The TriNetX platform is compliant with the Health Insurance Portability & Accountability Act and General Data Protection Regulation. The Western Institutional Review Board has granted TriNetX a waiver of informed consent as this platform only records counts and statistical summaries of de-identified patient information.

#### Cohort Definition and Eligibility

A total of 93,656,801 adults aged 18-80, male or female, were identified and included in the study. Participants with pre-existing Type 2 Diabetes Mellitus (ICD-10-CM E11) who underwent catheter-based ablation (CPT93654) procedure for ventricular tachycardia were made part of the study. This narrowed down the total participants to 28,418.

#### Baseline Characteristics

After the initial selection of participants, they were stratified into sodium– glucose cotransporter-2 inhibitors (SGLT2i) receivers (n=18,652) and non-receivers (n=9,766) at the time of undergoing ablation. Given the significant differences in baseline characteristics between participants, we included the following covariate factors: demographics (age, gender, ethnicity), underlying conditions such as Heart failure (ICD-10-CM I50),

Cardiomyopathy (ICD-10-CM I42), Long QT syndrome (ICD-10-CM 145.81), Hypertensive diseases (ICD-10-CM I10-I1A), Ischemic heart diseases ( ICD-10-CM I20-I25), Disorders of lipoprotein metabolism and other lipidemia (ICD-10-CM E78), Hypokalemia (ICD-10-CM E87.6), Hyperkalemia (ICD-10-CM E87.5), Disorders of magnesium metabolism (ICD-10-CM E83.4), Disorders of calcium metabolism (ICD-10-CM E83.5), Disorders of thyroid gland (ICD-10-CM E00-E07), Chronic kidney disease, stage 3 (ICD-10-CM N18.3), Chronic kidney disease, stage 4 (ICD-10-CM N18.4), Chronic kidney disease, stage 5 (ICD-10-CM N18.5), BMI 30-39 (ICD-10-CM Z68.3), BMI 40 or greater (ICD-10-CM Z68.4). Moreover, we included potential confounding factors such baseline medication use, laboratory results, corrected QT interval, and left ventricular ejection fraction (LVEF) as seen in the flowchart in Figure 1.

**Fig 1:**
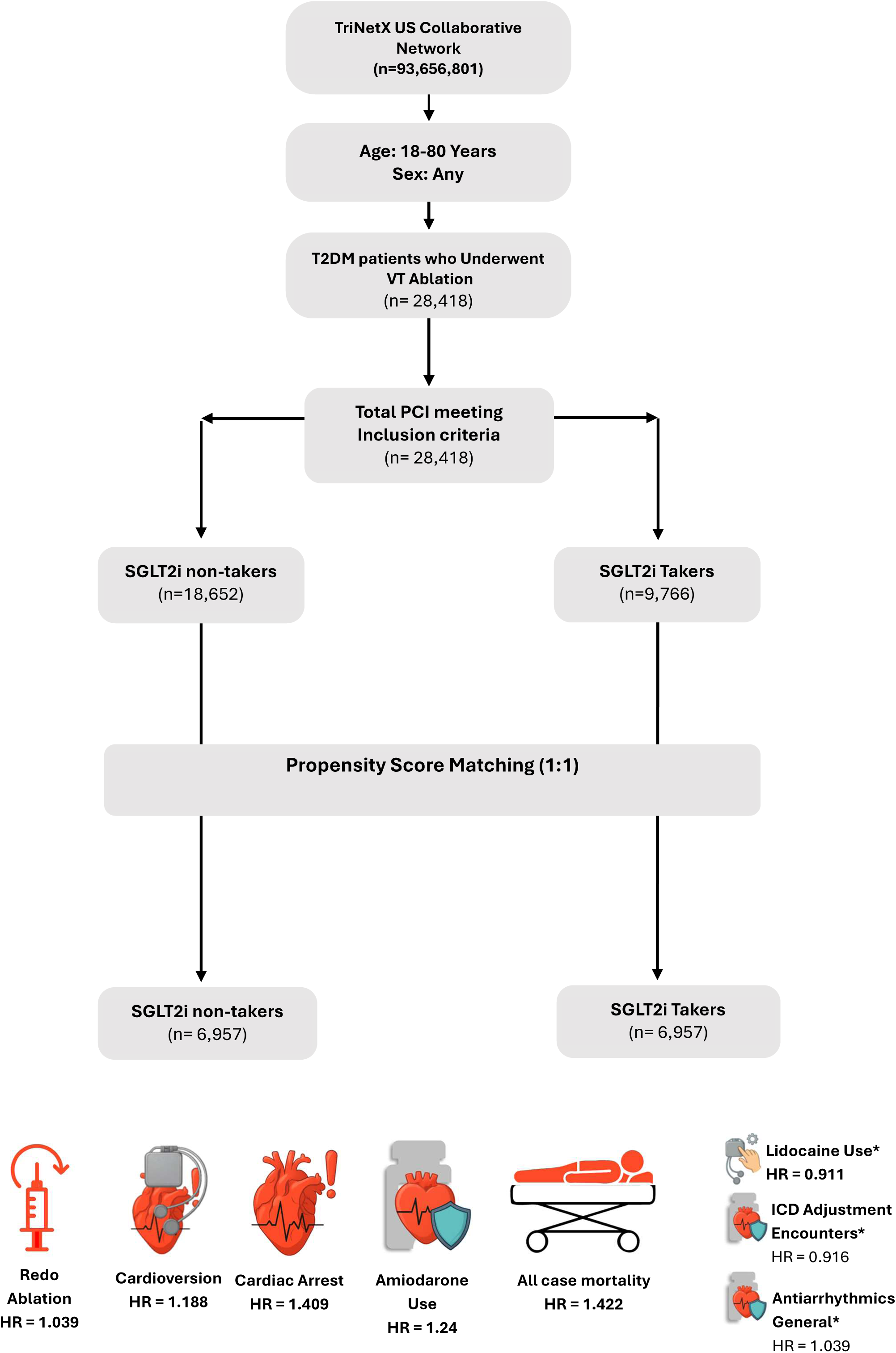
Flow Diagram of patient selection. From the TriNetX US collaborative Network (n 93,656,801), patients aged 18-80 years with diabetes type II who underwent ventricular tachycardia ablation were identified (n=28,418). Of these 18,652 were non-users and 9,766 were users of SGLT2i. Following propensity score matching for demographic, comorbid, and laboratory variables 6,957 patients remained in each cohort. HR indicates Hazard Ratio with asterisk mark [*] indicating non-significance.

### Outcomes and Endpoints

The initial ablation was identified as the index ablation procedure, and patients were followed for 3 years post-ablation to assess the primary and secondary outcomes. Primary outcomes were characterized as all-cause mortality and occurrences of cardiac arrest after the index ablation within the time. Secondary outcomes were defined as visits for ICD adjustments, post ablation cardioversion, post ablation utilization of lidocaine, amiodarone, or any class of antiarrhythmics.

### Statistical Analysis

We utilized propensity score matching to minimize the effect of confounding factors and divide participants into groups with matched baseline characteristics. By using TriNetX built-in function, we matched the two groups at a 1:1 ratio by greedy nearest neighbor algorithm. Factors considered included demographics such as age, sex, ethnicity, and underlying comorbid conditions. We also considered baseline medication use, and laboratory values. The Hazard ratio (HR), odds ratio (OR), risk ratio (RR), and risk difference (RD) were calculated for primary and secondary outcomes after VT ablation in SGLT2i non-users versus users after propensity score matching. Outcomes were assessed within 3 years following the index ablation procedure. Patients with a recorded occurrence of the primary or secondary outcomes prior to the index ablation event were excluded from each respective outcome analysis. We employed the Kaplan-Meier method log-rank tests for the survival probability analysis. In all our analyses, a 95% confidence interval (95% CI) was considered evidence of statistical significance, with the P-value < 0.05.

## Results

### Study population

A total of 28,418 patients (aged 18 to 80 years) with type 2 diabetes mellitus (T2DM) who underwent catheter ablation for ventricular tachycardia (VT) were identified from the TriNetX US Collaborative Network. At the time of ablation, 9,766 (34.4%) were receiving sodium–glucose cotransporter-2 inhibitors (SGLT2i), and 18,652 (65.6%) were not. Prior to propensity score matching, there were significant imbalances in baseline demographics, comorbidities, medication use, and laboratory parameters between the two groups. (Table 1. Basic Characteristics Table)

**Table 1:** Baseline characteristics of cohorts before and after propensity matching. Numerical data is shown as mean ± standard deviation, whereas categorical data is represented as count (%). Abbreviations: SD: Standard Deviation, Std diff: Standard Difference, BMI: Body mass index.

| Variables | SGLT2i non-Takers | SGLT2i Takers | P-Value | Std diff. |
| --- | --- | --- | --- | --- |
| **Demographics** |  |  |  |  |
| Current Age | 6,957 | 6,957 | 0.428 | 0.013 |
| White | 5,027 | 5,051 | 0.649 | 0.008 |
| Female | 1,970 | 1,962 | 0.880 | 0.003 |
| Unknown Ethnicity | 1,253 | 1,248 | 0.912 | 0.002 |
| Not Hispanic or Latino | 5,310 | 5,328 | 0.719 | 0.006 |
| Hispanic or Latino | 394 | 381 | 0.631 | 0.008 |
| Black or African American | 820 | 811 | 0.813 | 0.004 |
| Male | 4,659 | 4,671 | 0.829 | 0.004 |
| **Diagnosis** |  |  |  |  |
| Heart failure | 4,403 | 4,404 | 0.986 | <0.001 |
| Cardiomyopathy | 2,855 | 2,855 | 1 | <0.001 |
| Long QT syndrome | 491 | 501 | 0.742 | 0.006 |
| Hypertensive diseases | 6,368 | 6,375 | 0.831 | 0.004 |
| Ischemic heart diseases | 4,508 | 4,533 | 0.657 | 0.008 |
| Disorders of lipoprotein metabolism and other lipidemia | 5,622 | 5,620 | 0.966 | 0.001 |
| Hypokalemia | 1,414 | 1,411 | 0.950 | 0.001 |
| Hyperkalemia | 925 | 935 | 0.803 | 0.004 |
| Disorders of magnesium metabolism | 942 | 947 | 0.902 | 0.002 |
| Disorders of calcium metabolism | 502 | 509 | 0.819 | 0.004 |
| Disorders of thyroid gland | 1,880 | 1,837 | 0.410 | 0.014 |
| Chronic kidney disease, stage 3 (moderate) | 1,183 | 1,203 | 0.653 | 0.008 |
| Chronic kidney disease, stage 4 (severe) | 360 | 377 | 0.520 | 0.011 |
| Chronic kidney disease, stage 5 | 93 | 99 | 0.663 | 0.007 |
| Body mass index [BMI] 30-39, adult | 2,342 | 2,360 | 0.747 | 0.005 |
| Body mass index [BMI] 40 or greater, adult | 1,277 | 1,257 | 0.660 | 0.007 |
| **Medication** |  |  |  |  |
| Beta blockers/related | 6,511 | 6,507 | 0.890 | 0.002 |
| Antiarrhythmics | 6,304 | 6,265 | 0.263 | 0.019 |
| Atorvastatin | 4,483 | 4,442 | 0.469 | 0.012 |
| Rosuvastatin | 1,540 | 1,547 | 0.886 | 0.002 |
| Simvastatin | 1,158 | 1,130 | 0.522 | 0.011 |
| Pravastatin | 982 | 978 | 0.922 | 0.002 |
| Lovastatin | 135 | 140 | 0.761 | 0.005 |
| Pitavastatin | 29 | 29 | 1 | <0.001 |
| Diuretics | 5,549 | 5,559 | 0.833 | 0.004 |
| Calcium channel blockers | 4,504 | 4,502 | 0.972 | 0.001 |
| Ace inhibitors | 3,576 | 3,580 | 0.946 | 0.001 |
| Angiotensin ii inhibitor | 3,680 | 3,667 | 0.825 | 0.004 |
| Digitalis glycosides | 1,306 | 1,310 | 0.931 | 0.001 |
| Anticoagulants | 6,647 | 6,645 | 0.935 | 0.001 |
| Platelet aggregation inhibitors | 5,027 | 5,030 | 0.955 | 0.001 |
| Thyroid supplements | 1,281 | 1,228 | 0.243 | 0.020 |
| Sympathomimetics (adrenergic) | 4,106 | 4,095 | 0.850 | 0.003 |
| **Laboratory** |  |  |  |  |
| Potassium [Moles/volume] in Serum, Plasma or Blood | 6,361 | 6,388 | 0.952 | 0.001 |
| Calcium [Mass/volume] in Serum, Plasma or Blood | 6,386 | 6,387 | 0.095 | 0.030 |
| Magnesium [Mass/volume] in Serum, Plasma or Blood | 4,849 | 4,837 | 0.350 | 0.019 |
| Troponin I,cardiac [Mass/volume] in Serum, Plasma or Blood | 2,307 | 2,270 | 0.125 | 0.045 |
| Natriuretic peptide B [Mass/volume] in Serum, Plasma or Blood | 2,665 | 2,691 | 0.034 | 0.058 |
| Natriuretic peptide. B prohormone N-Terminal [Mass/volume] in Serum, Plasma or Blood | 1,939 | 2,035 | 0.001 | 0.105 |
| Hemoglobin A1c/Hemoglobin. Total in Blood | 5,296 | 5,128 | <0.001 | 0.144 |
| Corrected QT Interval (QTc) | 1,178 | 1,054 | 0.057 | 0.081 |
| Left Ventricular Ejection Fraction (LVEF) (%) | 1,426 | 1,402 | 0.249 | 0.043 |

**Table 2:** Cohort 1 and cohort 2 patient count before and after propensity score matching.

| Cohort | Patient count before matching | Patient count after matching |
| --- | --- | --- |
| SGLT<sub>2</sub>i non-Takers | 18,652 | 6,957 |
| SGLT<sub>2</sub>i Takers | 9,766 | 6,957 |

**Fig 2:**
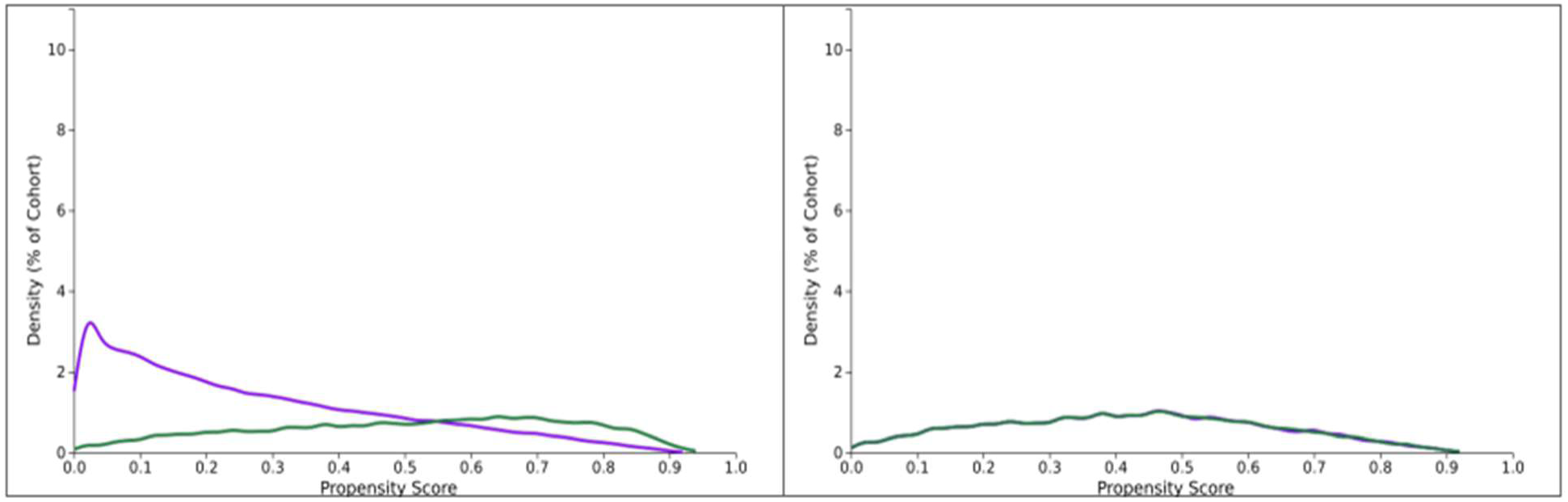
Propensity score density function - Before and after matching (cohort 1 - purple, cohort 2 - green). This figure compares propensity score distributions for two cohorts before and after matching. After matching (right-sided figure) the distributions overlap significantly, confirming improved balance between both the cohorts.

Following 1:1 propensity score matching, 6,957 patients remained in each cohort. Matching achieved excellent covariate balance, with the standardized mean differences reduced to <0.05 for nearly all variables, and no statistically significant differences observed between groups in demographics, comorbidities, medication use, or laboratory values. (Table 1. Basic Characteristics Table)

### Primary outcomes

Over a median follow-up of 3 years after VT ablation, SGLT2 inhibitor non-users exhibited significantly higher hazard ratios (HR) for various adverse outcomes. (Table 3: Outcomes Table)

**Table 3:**
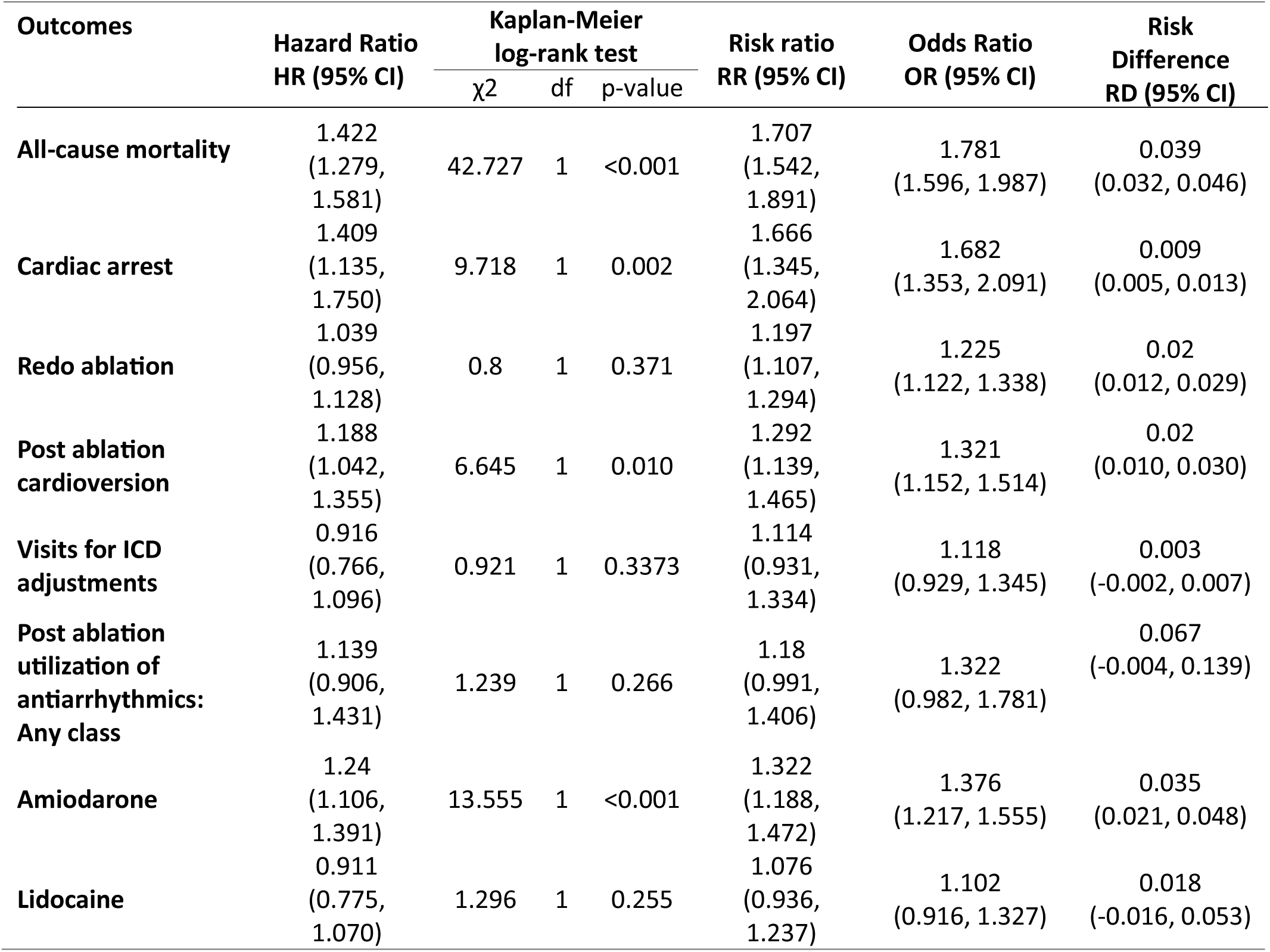
Hazard ratio, odds ratio, risk ratio, and risk difference for outcomes after VT ablation in SGLT2i non-users versus users after propensity score matching. Abbreviations/Key: CI = Confidence interval; df = Degrees of freedom; HR = Hazard ratio; RD = Risk difference; RR = Risk ratio; OR = Odds ratio; VT = Ventricular tachycardia; SGLT2i = Sodium glucose linked transporter inhibitor; ICD = Implantable cardioverter-defibrillator.

All-cause mortality occurred in 931 non-users (13.4%) and 545 users (7.8%), corresponding to a hazard ratio (HR) of 1.422 (95% CI: 1.279-1.581; p < 0.001). Cardiac arrest was observed in 219 non-users (3.1%) and 132 users (1.9%), with an HR of 1.409 (95% CI: 1.135-1.750; p = 0.002).

### Secondary outcomes

Post-ablation cardioversion occurred more frequently in non-users (514 events; 7.4%) compared to users (396 events; 5.7%), with an HR of 1.188 (95% CI: 1.042-1.355; p = 0.010). Post-ablation amiodarone use was higher among non-users (682 events; 9.8%) compared with users (513 events; 7.4%), HR 1.240 (95% CI: 1.106-1.391; p < 0.001).

No significant differences were observed for redo ablation (HR: 1.039; 95% CI: 0.956-1.128; p = 0.371), ICD adjustment visits (HR 0.916; 95% CI: 0.766-1.069; p = 0.337), use of any antiarrhythmic (HR: 1.139; 95% CI: 0.906-1.431; p = 0.266), or post-ablation lidocaine (HR: 0.911; 95% CI: 0.775-1.070; p = 0.255) between the two groups.

**Figure 3:**
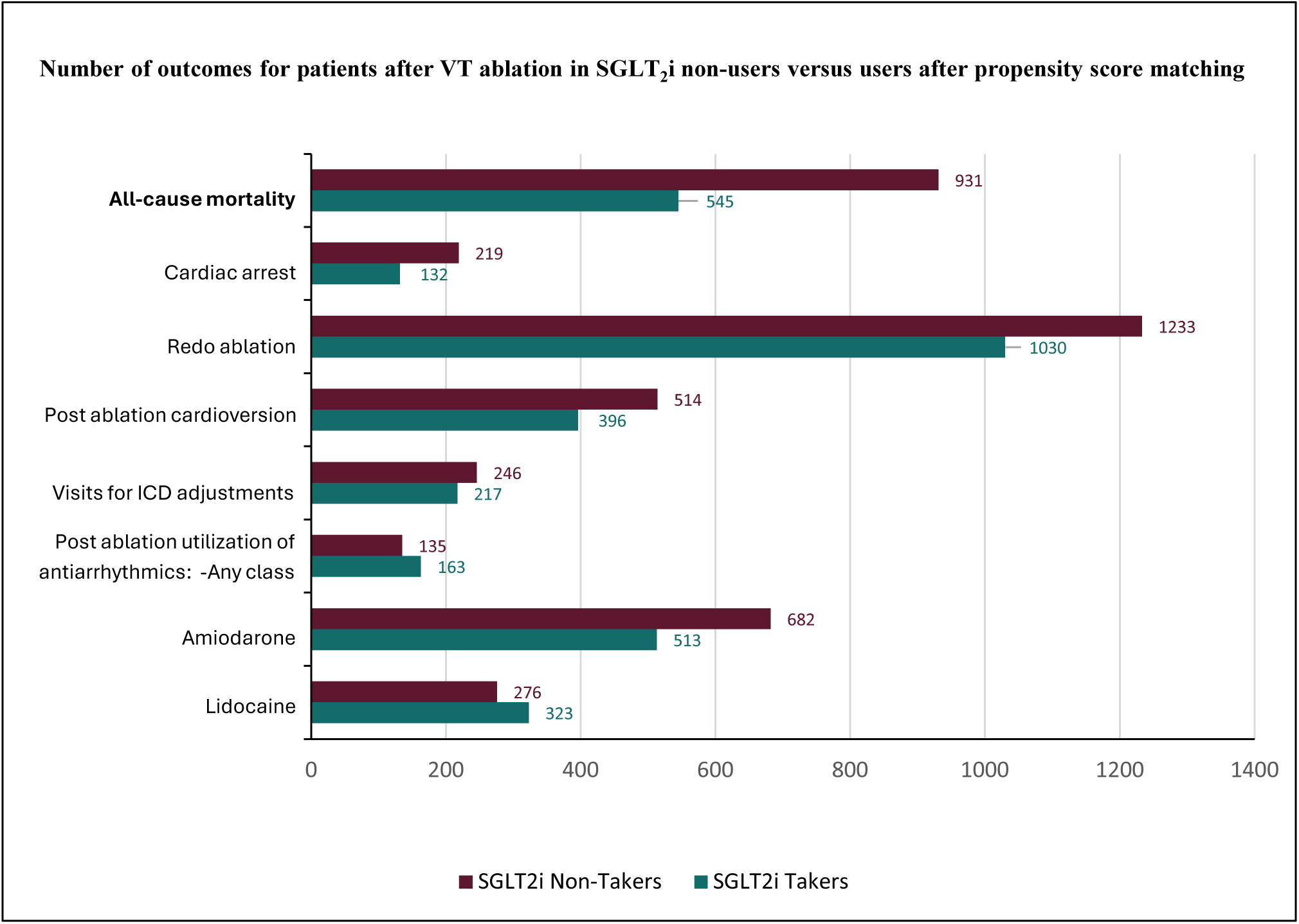
Bar chart comparing the number of outcomes for patients after VT ablation in SGLT2i non-takers vs takers

**Figure 4:**
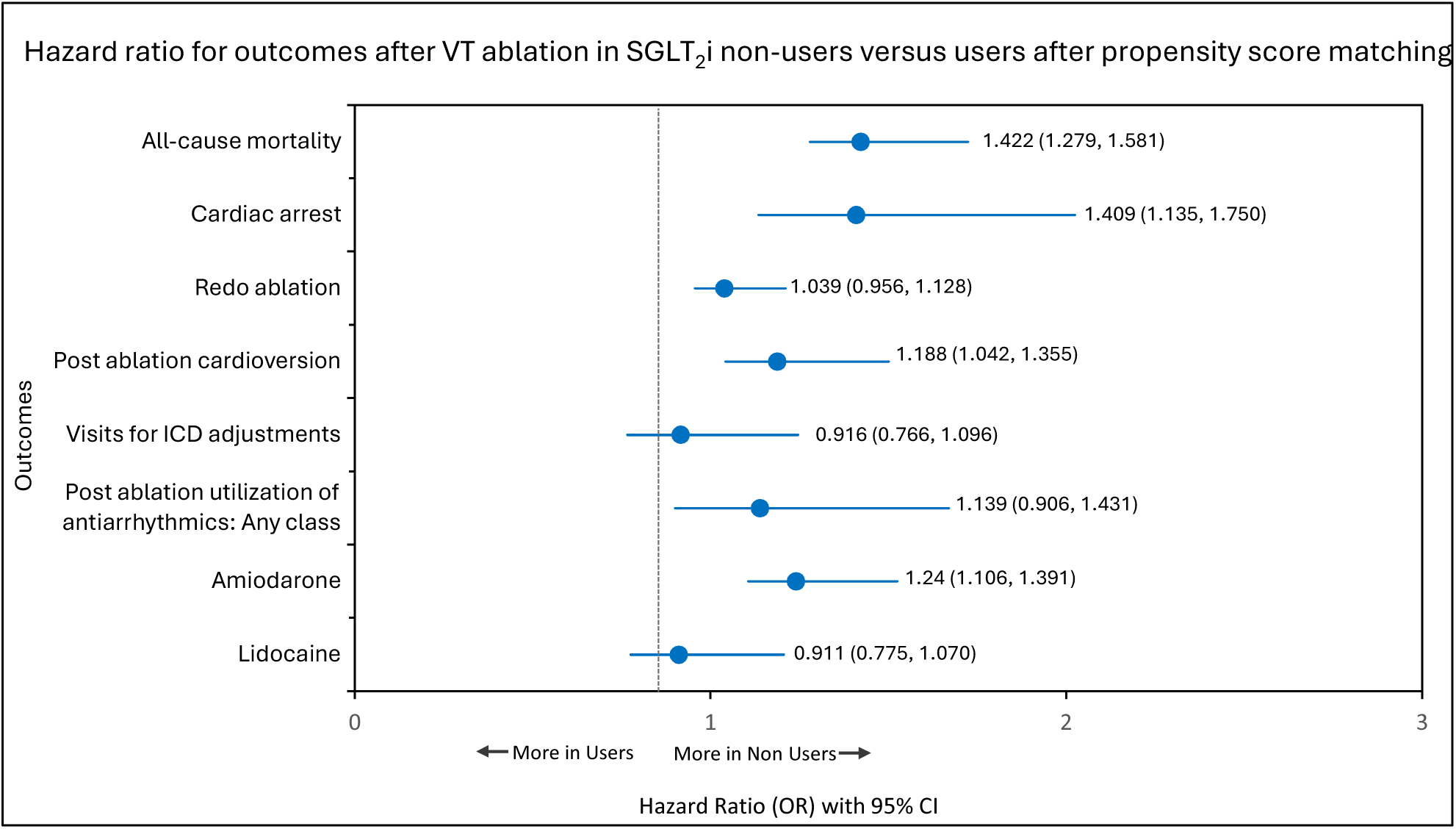
Forest plot: Hazard ratio with 95% CI for outcomes after ventricular tachycardia ablation in SGLT2i non-users versus users after propensity score matching

### Summary of effect estimates

The forest plot demonstrates that hazard ratios for all-cause mortality, cardiac arrest, post ablation cardioversion, and amiodarone use were consistently >1 and favored the SGLT2i user group, with 95% confidence intervals excluding unity. Other outcomes were centered near the null with wide confidence intervals, indicating no statistically significant association.

### Interpretation

In this large, propensity score–matched analysis of patients with T2DM undergoing VT ablation, SGLT2i use was associated with substantial reductions in all-cause mortality, cardiac arrest, and certain post-procedural interventions. These findings suggest a potential cardioprotective role of SGLT2i in the peri- and post-ablation setting. The effect was not observed for procedural recurrence metrics (redo ablation) or device-related outcomes (ICD adjustments), suggesting the cardioprotective effect may be endpoint-specific.

## Discussion

In this large, multicenter, real-world analysis using the TriNetX network, peri-procedural use of sodium–glucose cotransporter 2 inhibitors (SGLT2i) among patients undergoing ventricular tachycardia (VT) ablation were linked to markedly lower rates of all-cause mortality, cardiac arrest, and repeat ablation compared with matched non-users. After propensity score matching to balance demographic characteristics, comorbidities, baseline medications, and laboratory profiles, SGLT2i therapy remained independently associated with favorable post-ablation outcomes, including a reduced need initiating amiodarone following the procedure.

The cardiovascular benefits of SGLT2i extend beyond glycemic control, with robust evidence from large randomized controlled trials. In EMPA-REG OUTCOME, empagliflozin reduced cardiovascular death by 38%, hospitalization for heart failure by 35%, and all-cause mortality by 32% in patients with type 2 diabetes and established cardiovascular disease.(15) In the DAPA-HF trial, dapagliflozin reduced the risk of worsening heart failure or cardiovascular death by 26% in patients with reduced ejection fraction, irrespective of diabetes status. (16) Similarly, EMPEROR-Reduced demonstrated a 25% relative risk reduction in the composite of cardiovascular death or heart failure hospitalization with empagliflozin in patients with symptomatic HFrEF. (17) The consistency of these findings in high-risk cardiovascular populations aligns with our study and increases the possibility that SGLT2i therapy could confer similar mortality and morbidity benefits in patients undergoing VT ablation. Beyond their well-established benefits in heart failure, SGLT2i may also exert direct anti-arrhythmic effects. Bonora et al. reviewed preclinical and clinical data suggesting SGLT2i may reduce atrial and ventricular arrhythmia incidence through improved autonomic tone, attenuation of myocardial fibrosis, and modulation of ion channel activity. (18) In a murine model of diabetic cardiomyopathy, Shao et al. demonstrated that empagliflozin reduced the burden of ventricular arrhythmias, helped stabilize action potentials, and lessened myocardial fibrosis. (19)In a post-hoc analysis of EMPA-REG OUTCOME, Böhm et al. reported a modest reduction in resting heart rate and fewer investigator-reported arrhythmia events among empagliflozin users, suggesting a potential rhythm-stabilizing effect in addition to its established benefits on heart failure and survival.(20) These results provide clinical backing for our findings.

In our real-world VT ablation cohort, patients who were on SGLT2i around the time of the procedure had fewer life-threatening arrhythmias and were less likely to need another ablation, adding weight to the idea that these drugs could play a protective, anti-arrhythmic role in high-risk patients. Several biological mechanisms may underlie the observed associations. SGLT2i promote osmotic diuresis and natriuresis, leading to reduced ventricular filling pressures and improved hemodynamic stability. SGLT2i promote osmotic diuresis, which reduces intravascular volume and alleviates congestion. (21) They also induce natriuresis, lowering ventricular filling pressures and helping to optimize preload. (22) Additionally, attenuates myocardial fibrosis, potentially decreasing arrhythmogenic substrate, improve myocardial energetics by shifting substrate utilization toward ketone metabolism.(23) Moreover, reduces inflammation and oxidative stress, further supporting myocardial recovery and stability. (24) These effects may collectively decrease arrhythmogenic substrate and improve left ventricular ejection fraction, thereby lowering the risk of VT recurrence and need for repeat interventions. The reduced amiodarone use observed in our analysis may reflect decreased arrhythmia burden in the SGLT2i group.

Due to the high morbidity and mortality associated with recurrent VT after ablation, strategies that improve long-term procedural success are clinically important. Our findings suggest that initiating or continuing SGLT2i therapy in eligible patients undergoing VT ablation could be a valuable adjunctive measure. (25) While causality cannot be inferred from observational data, the magnitude and consistency of risk reduction across multiple clinically relevant endpoints underscore the potential importance of integrating SGLT2i into post-ablation management algorithms for patients with concomitant heart failure or diabetes. The reduction in arrhythmia burden observed in our study is consistent with findings from device-based monitoring cohorts, where SGLT2i initiation led to fewer atrial and ventricular arrhythmia episodes.(26)

One of the key strengths of this study is its large, geographically diverse patient pool, drawn from multiple institutions, which adds to the generalizability of the findings. The use of rigorous propensity score matching further helped reduce the impact of confounding factors.(27) Nevertheless, several limitations warrant consideration. First, the retrospective nature of the study precludes definitive causal inference. Residual confounding from unmeasured variables—such as left ventricular scar burden, procedural techniques, or medication adherence—remains possible. Second, reliance on ICD and CPT codes may introduce misclassification, as coding variations in documentation practices can affect diagnostic accuracy.(28) Third, TriNetX provides de-identified, aggregate-level data, which limits the ability to assess arrhythmia mechanisms, procedural complications, or cause-specific mortality. (29) Finally, the findings may not be generalizable to populations outside the contributing health systems.

Looking ahead, prospective randomized trials are needed to confirm whether SGLT2i use truly protects patients undergoing VT ablation and to better understand the mechanisms involved. (30) It would also be valuable to explore whether these benefits apply to patients without diabetes, across diverse cardiomyopathy etiologies, and in combination with other anti-arrhythmic or neurohormonal therapies. (23)

## Data Availability

All data produced are available online at https://live.trinetx.com/

## Statements and Declarations

### Ethical Approval

No ethical approval was required for the study

### Consent

No consent was needed

## Acknowledgments

None

## Data Availability Statement

All data generated or analyzed during this study are included in this article. Further inquiries can be directed to the corresponding author.

